# MRI2PET: Realistic PET Image Synthesis from MRI for Automated Inference of Brain Atrophy and Alzheimer’s

**DOI:** 10.1101/2025.04.23.25326302

**Authors:** Brandon Theodorou, Anant Dadu, Brian Avants, Mike Nalls, Jimeng Sun, Faraz Faghri

**Affiliations:** Department of Computer Science, University of Illinois at Urbana-Champaign, Urbana, IL, USA; Center for Alzheimer’s and Related Dementias, National Institutes of Health, Bethesda, MD, USA; Laboratory of Neurogenetics, National Institute on Aging, National Institutes of Health, Bethesda, MD, USA; DataTecnica, LLC, Washington, DC, USA; University of Virginia, Dept of Radiology and Medical Imaging, Charlottesville, VA, USA

## Abstract

**Background:** Positron Emission Tomography (PET) scans are a crucial tool in the diagnosing and monitoring of a number of complex conditions, including cancer, heart health, and especially cognitive brain function. However, they are also often much more expensive than comparable imaging modalities such as X-Ray and magnetic resonance imaging (MRI), which can limit their availability and the impact of their use in both medical and machine learning settings. We propose to address this problem by using generative models to simulate the PET scan results based on prior MRI.

**Methods:** While recent work has yielded impressive realism in image generation, this PET synthesis task presents a series of technical challenges based on the scarcity of paired data as well as the complexity and nuance of the 3D images. So, we propose MRI2PET to generate AV45-PET scans from T1-weighted MRI images. MRI2PET is a 3D diffusion-based method which makes use of style transferred pre-training and a Laplacian pyramid loss to address these challenges by utilizing larger available unpaired MRI datasets and structural similarities between the MRI and PET images while simultaneously emphasizing the crucial details.

**Findings:** We evaluate MRI2PET through a series of studies on the ADNI dataset where we show that it both generates realistic images and improves clinically-based disease classification. When compared to training on only the original AV45-PET data, MRI2PET augmentation increases AUROC of brain scan classification to 0.780 ± 0.005 from 0.688 ± 0.014 when classifying brain scans into one of three clinically defined groups: cognitively normal, mild cognitive impairment, and Alzheimer’s Disease.

**Interpretation:** The capability to generate high quality, clinically relevant PET scans from MRI has the potential to expand the utility of cost-effective and accessible imaging workflows and improve both image-based machine learning capabilities and patient care.

**Funding:** US National Institute on Aging, US National Institutes of Health, US National Science Foundation.

## Introduction

Positron Emission Tomography (PET) scans represent a cornerstone of advanced medical imaging diagnostics. These molecular imaging studies provide crucial insights into metabolic processes,^13^ enabling precise detection of cancer,^12,27,41^ detailed evaluation of brain function^23^, and comprehensive assessment of cardiac health.^7,26^ Specifically, their ability to provide nuanced clinical details, such as precise tumor metabolic activity, makes them invaluable for personalized treatment planning.^33,40^ However, the widespread clinical utilization of PET imaging faces significant barriers, including high operational costs, limited availability of radiopharmaceuticals, and restricted access to specialized facilities.^18,19^ These limitations not only constrain the medical value of PET imaging but also severely restrict the development of machine learning models, as PET datasets remain substantially smaller compared to other, often less specific but more broadly available, imaging modalities like Magnetic Resonance Imaging (MRI). Finding innovative approaches to make the benefits of PET scans more accessible, even in a simulated capacity, would significantly advance early diagnosis capabilities, enhance treatment planning protocols, and enable the development of more robust AI-based clinical tools.

Recent advances in deep learning, particularly in generative models, offer a promising avenue for simulating PET scans from readily available data. The image generation domain has grown rapidly in recent years to the point of exceptional photorealism. This rise began largely with the introduction of Generative Adversarial Networks (GANs).^5,6,14^ GANs use adversarial training between a generator which creates data indistinguishable from real samples against a discriminator which tries to differentiate between real and generated data. GANs have gained widespread popularity for their ability to turn this simple training trick into incredibly realistic images. However, they have run into a number of challenges such as mode collapse and training instability.

So, a more direct approach which has re-popularized older score-based models to impressive success is the use of diffusion models.^8,34,35,36^ Diffusion models iteratively refine an image out of random noise through a series of small adjustments. They are a probabilistic model which has shown the ability to produce high-quality and complex images. Furthermore, they have shown to scale effectively, with largely such models yielding incredibly realistic results.^30,31^ Many of these diffusion model approaches have even generated conditioned images^3,30,38^ and 3D outputs ^9,10,11,29^ that can offer insights into our task at hand.

GANs and diffusion models have also demonstrated remarkable capabilities in medical image generation.^1,5,6,8,14,15,16,28,34,35,36^ Notably, conditional generation, where images are synthesized based on auxiliary information such as text or other imaging modalities, holds particular relevance.^17,39,42^ Given that PET scans are typically part of a comprehensive diagnostic imaging protocol, often following or complementing structural imaging modalities such as MRI to provide additional functional and metabolic information,^4,21^ leveraging MRI as a conditioning input for PET synthesis is a logical and cost-effective approach. Potentially democratizing access to PET-like diagnostic information without incurring the substantial costs and logistical challenges associated with actual PET acquisition.

However, generating accurate and clinically relevant PET images from MRI presents several technical challenges that must be systematically addressed:

- **Data Scarcity**. The limited availability of paired MRI-PET data compared to the abundance of MRI-only data presents a fundamental constraint that necessitates techniques that can effectively utilize unpaired data.
- **3D Complexity**. Unlike conventional 2D image generation tasks, PET scans represent complex three-dimensional volumetric data with intricate spatial relationships that must be preserved throughout the generation process.
- **Structural Fidelity**. PET and MRI scans exhibit structural similarity but differ fundamentally in their functional content. Ensuring anatomical fidelity and accuracy without structural distortion during generation is crucial.

To address these challenges, we propose **MRI2PET**, a novel 3D diffusion-based method for generating PET images from MRI scans. Our approach incorporates two key innovations:

- **Style Transferred Pre-Training**. We leverage the vast amount of MRI-only data by implementing a style transfer technique that generates synthetic MRI-to-PET-like image pairs, enabling effective pre-training of the diffusion model. Style transfer is a generative technique leveraging image classification models which involves “transferring” the style of one image (e.g., color, texture) onto another while preserving the content structure. It was originally popularized in the domain of artistic image generation, but we apply it here to create “PET-like” MRI images for pre-training. This then allows the model to learn the fundamental structural relationships between MRI and PET, maximizing the utilization of available data resources while mitigating the constraints imposed by limited paired MRI-PET datasets.
- **Laplacian Pyramid Loss Component**. To enhance the fidelity and generation of important diagnostic details beyond general structural correspondence, we introduce a Laplacian pyramid loss function. This loss component ensures that the generated PET images accurately capture the multi-scale features present in real PET scans, improving diagnostic accuracy.

We evaluate MRI2PET using the publicly available Alzheimer’s Disease Neuroimaging Initiative (ADNI) dataset, specifically on the task of generating AV45 PET scans from T1-weighted MRI imaging. While the dataset contains various types of PET images such as FDG, Amyloid, and Tau PET scans, we opt for AV45 as the most prevalent type of PET imaging within the dataset and one which under the amyloid PET classification is known to provide value in the detection and study of Alzheimer’s disease as opposed to more cancer-focused modalities like FDG PET. Our comprehensive evaluation includes quantitative metrics, qualitative assessments, and clinical case studies, demonstrating that MRI2PET can effectively model PET scans. We further show that the generated images capture clinically relevant details, enabling their use for data augmentation and improving the performance of downstream machine learning tasks. This capability to generate high quality, clinically relevant PET scans from MRI has the potential to expand the utility of cost-effective, automated, and accessible imaging workflows and improve the quality, efficiency, and availability of important patient diagnostics. It offers the potential to provide important PET-based diagnostic findings to more people at an earlier date to allow for earlier diagnosis and more targeted testing where relevant without needing to wait for the severity of the risk or condition to warrant the cost.

## Methods

### MRI2PET Methodology

Our proposed MRI2PET method, illustrated in Figure 1, employs a conditional diffusion modeling framework enhanced by two critical innovations designed to address key challenges in synthesizing PET images from MRI scans: Style Transferred Pre-Training and Laplacian Pyramid Loss Component.

**Figure 1:**
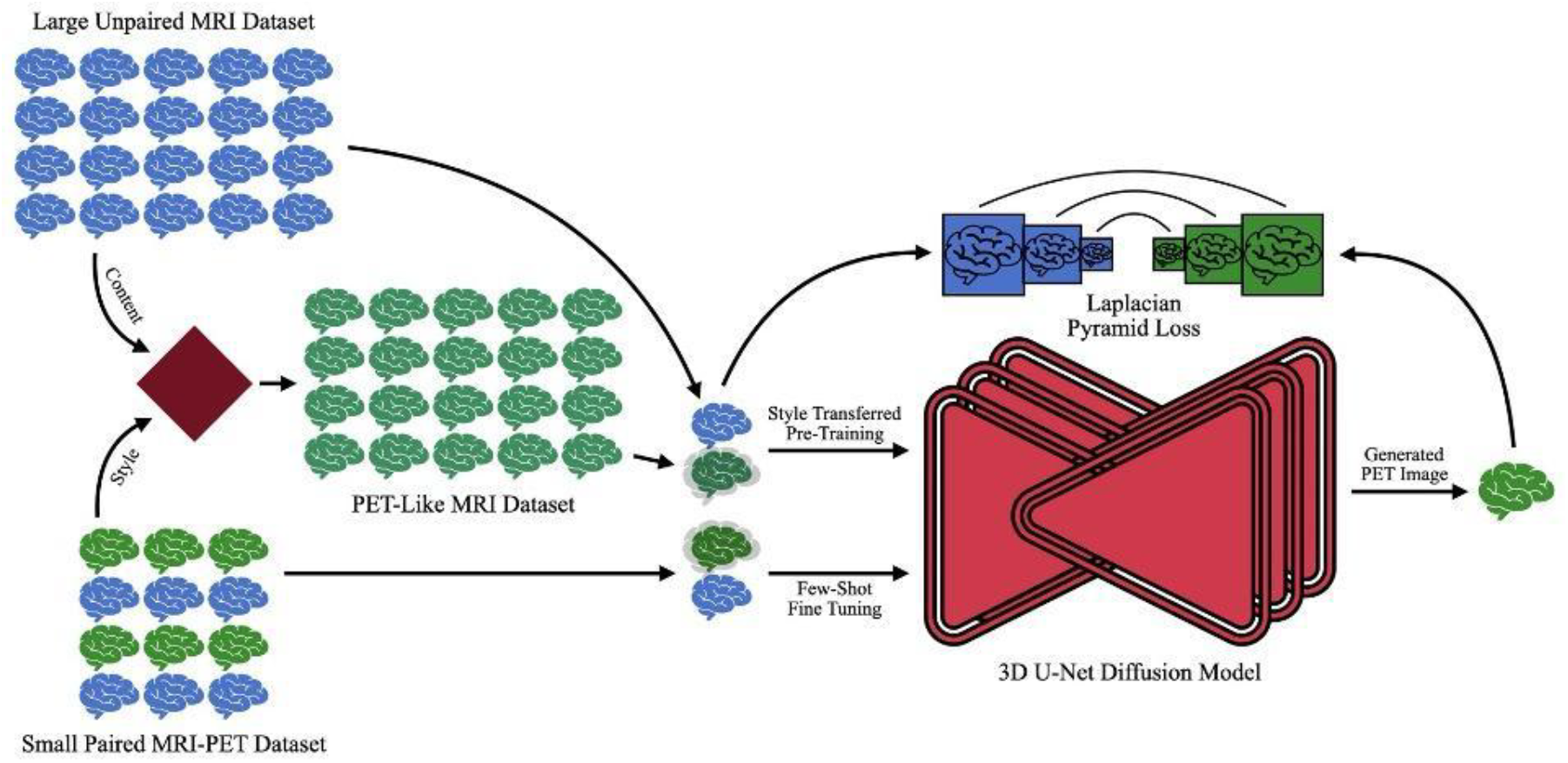
Schematic illustration of the MRI2PET architecture. The method employs a 3D U-Net diffusion model initially pre-trained on a large dataset of unpaired MRI images. Style transfer techniques are utilized to generate PET-like MRI images, effectively simulating MRI-to-PET conditions for pre-training. The model is subsequently fine-tuned on a smaller, paired MRI-PET dataset using a Laplacian pyramid loss, emphasizing the preservation and enhancement of critical multi-scale image details essential for clinical diagnostics.

#### Style Transferred Pre-Training

The primary challenge in generating clinically relevant PET images from MRI is the scarcity of paired MRI-PET datasets, due largely to the high cost and limited availability of PET imaging. To mitigate this data scarcity, we leverage abundant MRI-only datasets for pre-training. Rather than merely conditioning MRIs upon themselves (with added noise), we utilize a style transfer approach to closely simulate the PET imaging task. Specifically, a pre-trained VGG19 neural network is used to apply the style characteristics—such as intensity profiles and texture features—of real PET images to MRI slices. As VGG19 only processes 2D images, the style transfer is conducted slice-by-slice along the depth dimension of each MRI scan. This procedure generates synthetic MRI-to-PET-like image pairs, aligning pre-training more closely with the ultimate MRI-to-PET task. Consequently, the diffusion model is exposed early to critical PET-like features, significantly enhancing its ability to generate realistic PET images upon subsequent fine-tuning on limited paired datasets.

#### Laplacian Pyramid Loss Component

In addition to the conventional mean squared error (MSE)-based diffusion training objective (which instructs the model to predict the added noise), we introduce an auxiliary Laplacian pyramid loss to improve the model’s ability to capture fine diagnostic details at multiple resolutions. The Laplacian pyramid represents an image as a hierarchy of residuals between increasingly blurred (downsampled then upsampled) versions of the image, emphasizing details at varying scales. Specifically, this loss component is computed as follows:

- A synthetic PET image is generated from the diffusion model’s noise predictions combined with the input MRI scan.
- Five-level Laplacian pyramids are constructed separately for both synthetic and real PET images.
- At each pyramid level, the MSE between synthetic and real PET images is computed.
- MSE values across all pyramid levels are summed, forming the final Laplacian pyramid loss.

The addition of this loss encourages preservation of critical structural and textural details essential for more accurate image generation and clinical diagnostics. For more details see **Supplemental Material**.

### Dataset

We utilized publicly accessible paired MRI-PET scans from the Alzheimer’s Disease Neuroimaging Initiative (ADNI) dataset, which included 2,492 image pairs defined as scans from the same patient captured within one year.^22^Additionally, we incorporated 22,956 unpaired MRI images drawn from ADNI, UK Biobank,^37^ and Parkinson’s Progression Markers Initiative (PPMI) datasets.^20^ Tables of counts and demographic information of each of these datasets can be found in the supplemental material

All MRI scans underwent standard preprocessing steps, including template registration, skull stripping, and bias correction.^2^ MRI images were then resampled to a 64×144×144 voxel dimensionality using windowed sinc interpolation and normalized to a [0,1] intensity range.

PET images required handling temporal sequences, where applicable. Each frame was rigidly registered first to the initial frame, and subsequently to the temporal mean. The final PET image was the temporal mean, which was affine-registered to its corresponding MRI, resampled to a 32×128×128 voxel dimensionality via windowed sinc interpolation, and normalized.

For pre-training involving MRI outputs, MRI images were further resampled to PET dimensions (32×128×128) using quintic interpolation. We split the paired dataset randomly, reserving 20% for testing. The remaining paired data, combined with unpaired MRI scans, were further split into training (90%) and validation (10%) sets.

We pre-trained MRI2PET on the unpaired MRI dataset for 250 epochs, then fine-tuned on the paired MRI-PET dataset for 5,000 epochs. Training utilized the PyTorch framework,^25^ with a batch size of 128, learning rate of 0.0001, and the Adam optimizer.

### Baseline Comparisons

We benchmarked MRI2PET against external and internal baselines to validate performance comprehensively:

#### External Baselines

1. GAN: Conditional Wasserstein GAN trained on limited paired data.^6^
2. Vanilla Diffusion: Standard conditional diffusion model trained on limited paired data.
3. CDC: Few-shot GAN with cross-domain correspondences.^24^
4. DCL: Few-shot GAN optimizing diversity.^45^
5. DiffAugment: Differentiable augmentation GAN.^43^
6. MaskGAN: GAN with masked discriminator features.^46^
7. DDPM-PA: Few-shot diffusion model with pairwise adaptation.^47^

#### Ablation Studies

1. MRI2PET w/o Style Transfer: MRI-to-MRI noise-conditioned pre-training without style transfer.
2. MRI2PET w/o Pre-Training: Direct training without pre-training.
3. MRI2PET w/o Laplacian Loss: Training without additional Laplacian pyramid loss.

These rigorous comparative analyses were designed to highlight the contribution of each methodological innovation and validate MRI2PET’s overall effectiveness.

### Experimental Design

We rigorously evaluate MRI2PET using the publicly available ADNI dataset.^22^ Our experimental framework addresses three primary research questions:

1. Can MRI2PET generate realistic PET images from MRI?
2. Do synthetic PET images generated by MRI2PET replicate clinically meaningful patterns observed in actual patient PET images?
3. Does augmenting datasets with MRI2PET-generated PET images enhance performance in downstream machine learning tasks?

## Results

### MRI2PET Generates Realistic PET Images from MRI Scans

The primary aim of MRI2PET is the generation of realistic PET images from MRI data. We quantitatively assessed image quality using three established metrics: Fréchet Inception Distance (FID), Structural Similarity Index Measure (SSIM), and Peak Signal-to-Noise Ratio (PSNR). FID evaluates the similarity between distributions of real and synthetic images based on features extracted by a pre-trained Inception v3 model. Given that Inception v3 requires 2D inputs, we performed axial slicing of our 3D datasets. SSIM and PSNR were computed in 3D to evaluate structural detail preservation and noise levels, respectively. Results are summarized in Table 1 averaged over all of the images in our held-off test dataset.

**Table 1:** Quantitative evaluation of PET image generation quality comparing MRI2PET with various baseline and ablation methods. Metrics include Fréchet Inception Distance (FID), Structural Similarity Index Measure (SSIM), and Peak Signal-to-Noise Ratio (PSNR) calculated over the test set. Superior performance of MRI2PET across these metrics demonstrates enhanced realism, structural fidelity, and reduced noise in generated PET images. Note asterisks refer to p-value significance based on a two-tailed t-test comparing MRI2PET to the nearest non-ablation baseline with * p < 0.05, **p < 0.01, *** p < 0.001.

| Method | FID | SSIM | PSNR |
| --- | --- | --- | --- |
| GAN | 303.615 ± 0.063 | 0.101 ± 0.002 | 13.712 ± 0.025 |
| Vanilla Diffusion | 63.844 ± 0.046 | 0.764 ± 0.005 | 21.005 ± 0.137 |
| CDC | 287.531 ± 0.067 | 0.021 ± 0.000 | 7.780 ± 0.040 |
| DCL | 269.323 ± 0.045 | 0.076 ± 0.002 | 0.958 ± 0.015 |
| DiffAugment | 309.005 ± 0.052 | 0.055 ± 0.001 | 7.901 ± 0.028 |
| MaskGAN | 302.071 ± 0.048 | 0.016 ± 0.000 | 4.629 ± 0.020 |
| DDPM-PA | 265.000 ± 0.051 | 0.035 ± 0.000 | 8.108 ± 0.041 |
| w/o Style Transfer | 62.569 ± 0.025 | 0.854 ± 0.004 | 24.615 ± 0.130 |
| w/o Pre-Training | 108.401 ± 0.046 | 0.260 ± 0.008 | 15.644 ± 0.134 |
| w/o Loss | **61.213 ± 0.029** | 0.827 ± 0.004 | 23.156 ± 0.120 |
| MRI2PET | 61.735 ± 0.032*** | **0.857 ± 0.004\*\*\*** | **24.771 ± 0.123\*\*\*** |

MRI2PET significantly outperformed baseline methods across all metrics, particularly in SSIM and PSNR, indicating superior structural preservation, reduced noise, and enhanced visual quality in the generated PET images. While FID scores between MRI2PET and Vanilla Diffusion were similar, MRI2PET showed clear superiority over other baselines, with a notable improvement of more than 12% in SSIM and PSNR compared to the next-best external baseline. Ablation studies further clarified the roles of the individual contributions: removal of style-transferred pre-training significantly degraded performance, confirming its importance for effectively utilizing unpaired MRI data. Similarly, omitting Laplacian pyramid loss negatively impacted the quality of generated images, underscoring its crucial role in capturing detailed, multi-scale image features essential for clinical diagnostic accuracy. Collectively, these findings highlight the complementary strengths of these enhancements and their combined necessity for optimal performance.

Qualitative evaluations of axial slices from randomly selected test patients further reinforced these findings (Figure 2). MRI2PET consistently preserved patient-specific anatomical details such as brain size, shape, and orientation, closely mirroring real PET images and outperforming baselines.

**Figure 2:**
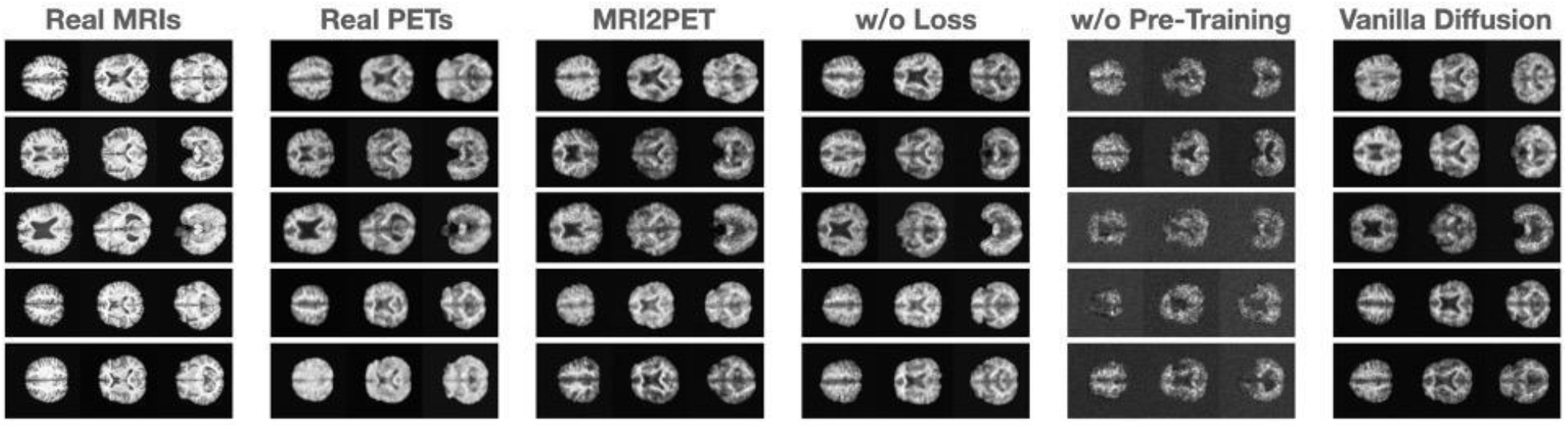
Qualitative comparison of axial brain slices from five randomly selected test patients. Images illustrate original MRI scans, real PET scans, and synthetic PET images generated by MRI2PET alongside baseline methods. MRI2PET notably maintains patient-specific anatomical structures, demonstrating high fidelity to the actual PET scans compared to other baselines.

Notably, structural intricacies, including distinct anatomical variations such as larger or smaller brain sizes and unique shape characteristics (e.g., horizontally elongated brains), were effectively captured. Moreover, MRI2PET reliably aligned physiological details between MRI and PET slices, demonstrating precise vertical alignment across modalities, which further validates the anatomical accuracy of generated images. These results provided a stark visual contrast compared to baseline methods, particularly highlighting MRI2PET’s substantial improvements over GAN-based and diffusion-based methods that consistently struggled with the complexities inherent in 3D image generation tasks.

### MRI2PET Captures Clinically Relevant Patterns

To examine MRI2PET’s ability to capture clinically meaningful patterns, we conducted detailed case-study analyses. In patients exhibiting significant brain atrophy (Figure 3), MRI2PET-generated PET images accurately reflected regions of decay observed in corresponding MRIs, demonstrating sensitivity to disease-specific anatomical changes. For instance, specific areas of notable atrophy, such as reduced cortical thickness or enlarged ventricles, were reliably replicated in the synthetic PET images, highlighting MRI2PET’s precision in capturing disease-associated morphological alterations. This speaks to the patient-specific and clinically relevant quality of MRI2PET’s generations, even as the atrophy itself may not be central to the downstream utility and usage of the PET imaging.

**Figure 3:**
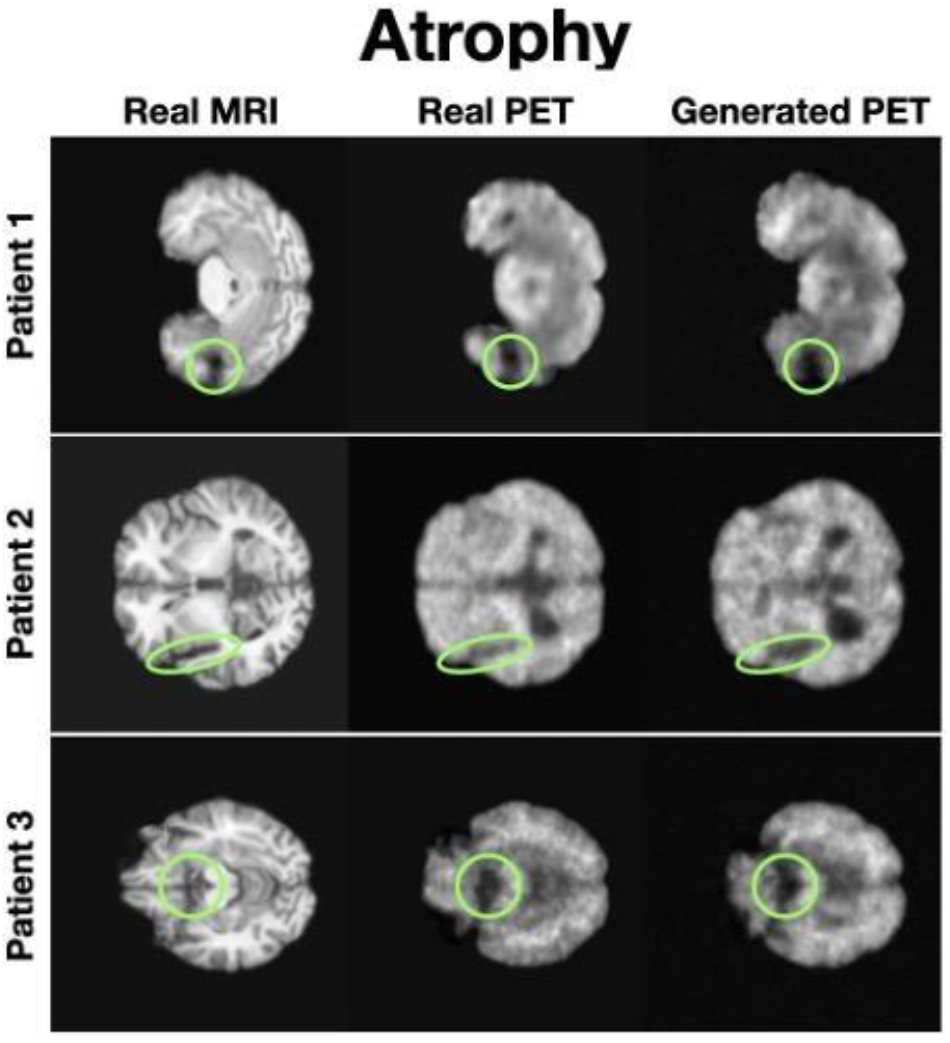
Case studies depicting detailed axial slices of three Alzheimer’s patients from the test set exhibiting severe brain atrophy. The figure presents a direct comparison between real MRI scans, actual PET scans, and MRI2PET-generated PET images, highlighting MRI2PET’s accurate capture and representation of disease-specific atrophy patterns and structural decay.

In longitudinal case studies involving Alzheimer’s disease progression (Figure 4), MRI2PET effectively captured deterioration patterns over time. Notably, increased prominence and depth of temporal sulci—a hallmark feature of progressive Alzheimer’s disease—were accurately reflected in sequential PET scans generated by MRI2PET. This temporal sensitivity to progressive anatomical degradation underscores MRI2PET’s potential utility for monitoring disease progression, facilitating timely interventions, and enabling more precise longitudinal clinical evaluations. These case studies collectively confirm MRI2PET’s robust capability in replicating disease-specific temporal and anatomical patterns, significantly contributing to its potential clinical utility in diagnostics and patient monitoring.

**Figure 4:**
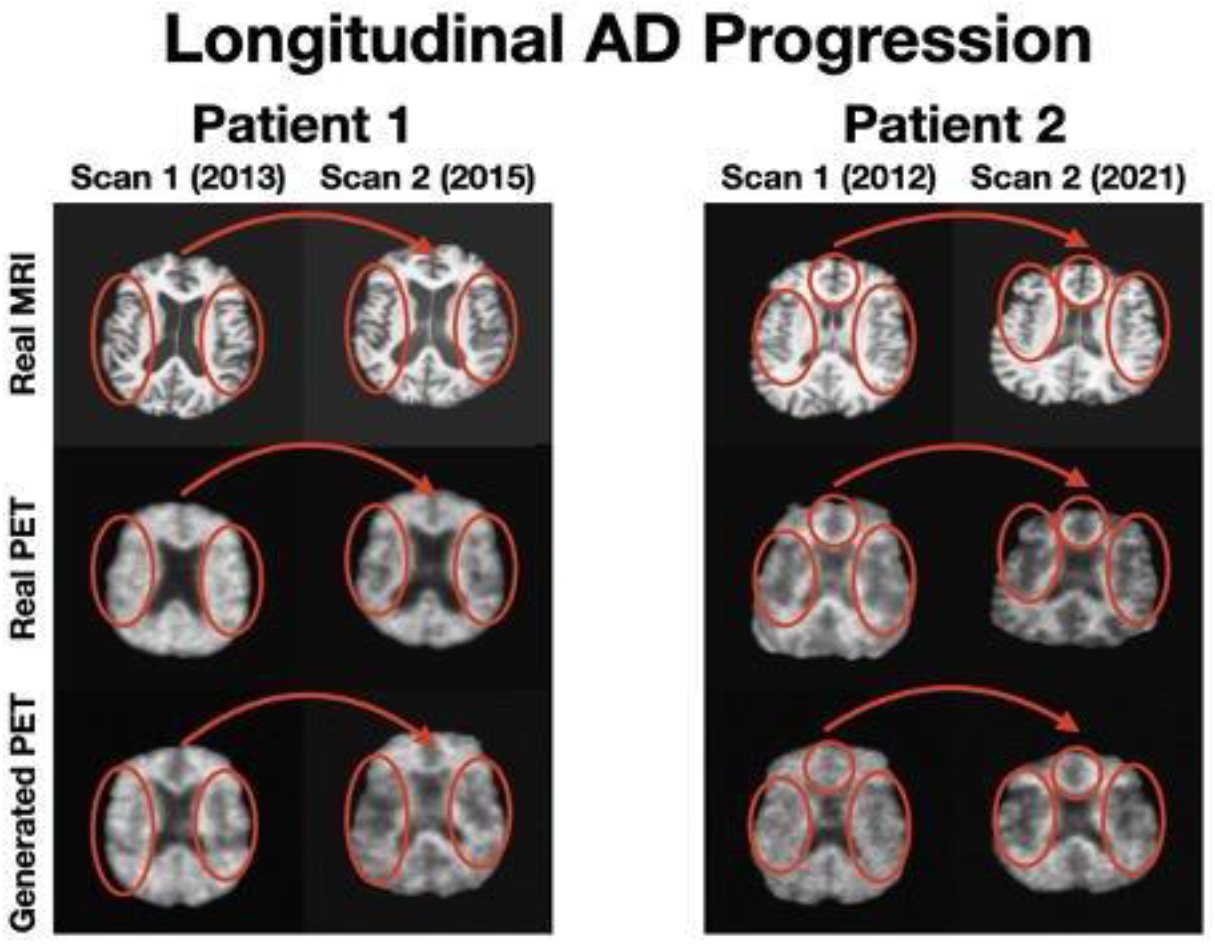
Additional detailed case studies involving two distinct test patients with two scans each illustrating longitudinal Alzheimer’s progression with increased brain atrophy in the second set of imaging. Side-by-side comparison of real MRI scans (top), real PET scans (middle), and corresponding synthetic PET scans generated by MRI2PET (bottom) clearly demonstrates MRI2PET’s capability to faithfully reproduce these patient-specific, clinically relevant details and structural variations even over time.

**Figure 5:**
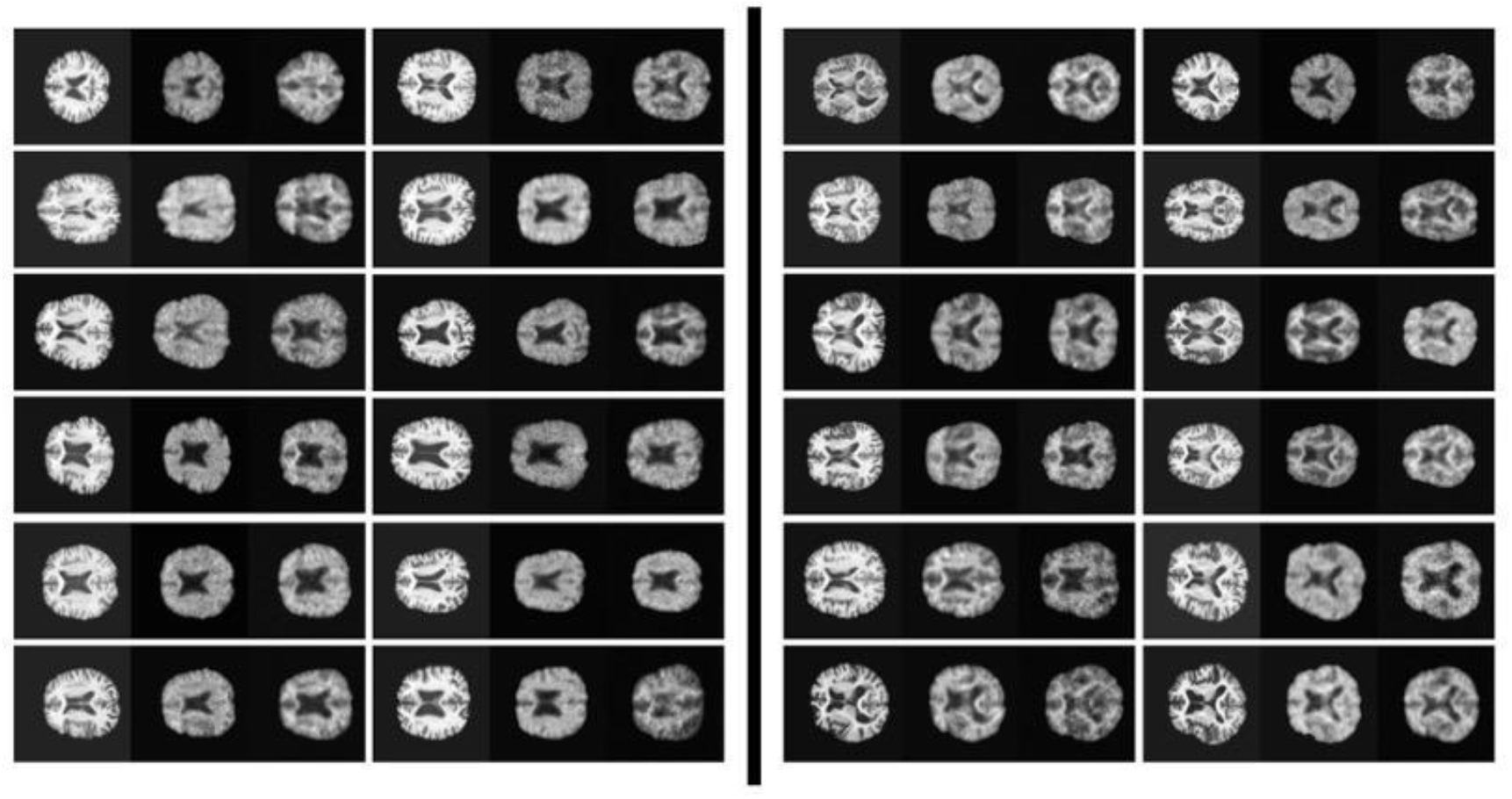
Comprehensive visualization of Alzheimer’s patient scans from the test dataset. Each image triplet includes the real MRI (left), real PET (middle), and corresponding MRI2PET-generated PET (right). Patients are broadly categorized, with those showing evident structural brain decay in MRI scans on the right half and those without visible decay on the left. Notably, generated PET images for patients without clear MRI-detectable decay generally exhibit greater blurriness, highlighting a correlation between generative uncertainty and early pathological stages, though exceptions occur on both sides.

### MRI2PET Enhances Downstream Machine Learning Tasks

We further assessed MRI2PET’s practical utility beyond visual realism by examining its impact on downstream machine learning tasks, specifically Alzheimer’s disease classification and Mini-Mental State Examination (MMSE) score prediction, leveraging clinically relevant patient labels from the ADNI dataset.

For Alzheimer’s disease classification, we utilized the ADNI dataset’s disease labels categorizing patients into cognitively normal, mildly cognitively impaired, or Alzheimer’s disease groups as a multi-class classification problem. The MMSE task employed scores from a standard cognitive examination, restricted to scores of 20 or above (out of a maximum of 30) to maintain focus on patients with mild or no cognitive impairment. Information regarding these MMSE distributions can be found in the demographics tables in the supplemental material.

We systematically evaluated three model types trained with distinct raw imaging inputs: MRI-only, PET-only, and MRI-PET paired data. Training involved multiple data configurations to comprehensively assess MRI2PET’s synthetic image utility:

- **Real MRI:** Large, unpaired MRI dataset.
- **Real Limited MRI:** Limited MRI data from paired MRI-PET scans.
- **Real PET:** Limited PET data without MRI pairing.
- **Synthetic PET:** Limited synthetic PET images generated by MRI2PET.
- **Real Paired:** Limited real MRI-PET pairs.
- **Augmented Paired:** Real MRI-PET pairs augmented by additional synthetic PET images paired with previously unpaired MRIs.

Performance across these datasets is detailed in Table 2 (classification) and Table 3 (MMSE regression) where we train 25 different models on the original training and validation datasets (the same as MRI2PET was trained on) using different random seeds for each task-model type pair and report the the mean metrics results along with the standard errors over those runs, calculated on the test dataset (which is paired and so can be used for any type of input).

**Table 2:** Performance results of downstream Alzheimer’s Disease classification tasks. The table compares test classification metrics (accuracy, F1 score, AUROC) across various data configurations of the original training dataset including real MRI, limited MRI, real PET, synthetic PET, and paired data (real and augmented). The augmented dataset incorporating MRI2PET-generated images on top of the limited existing real data significantly improves classification outcomes. Note asterisks refer to p-value significance based on a two-tailed t-test comparing augmented results to the nearest baseline approach with * p < 0.05, **p < 0.01, *** p < 0.001.

|  | Accuracy | F1 Score | Balanced Accuracy | AUROC |
| --- | --- | --- | --- | --- |
| Real MRI | $0.520 \pm 0.014$ | $0.497 \pm 0.016$ | $0.506 \pm 0.014$ | $0.704 \pm 0.012$ |
| Real, Limited MRI | $0.449 \pm 0.009$ | $0.369 \pm 0.020$ | $0.401 \pm 0.012$ | $0.611 \pm 0.014$ |
| Real PET | $0.542 \pm 0.009$ | $0.473 \pm 0.020$ | $0.498 \pm 0.013$ | $0.688 \pm 0.014$ |
| MRI2PET Generated PET | $0.542 \pm 0.009$ | $0.468 \pm 0.019$ | $0.496 \pm 0.012$ | $0.685 \pm 0.012$ |
| Real Paired | $0.559 \pm 0.006$ | $0.518 \pm 0.006$ | $0.526 \pm 0.007$ | $0.723 \pm 0.004$ |
| MRI2PET Augmented Paired | **$0.605 \pm 0.007^{***}$** | **$0.595 \pm 0.008^{***}$** | **$0.594 \pm 0.008^{***}$** | **$0.780 \pm 0.005^{***}$** |

**Table 3:**
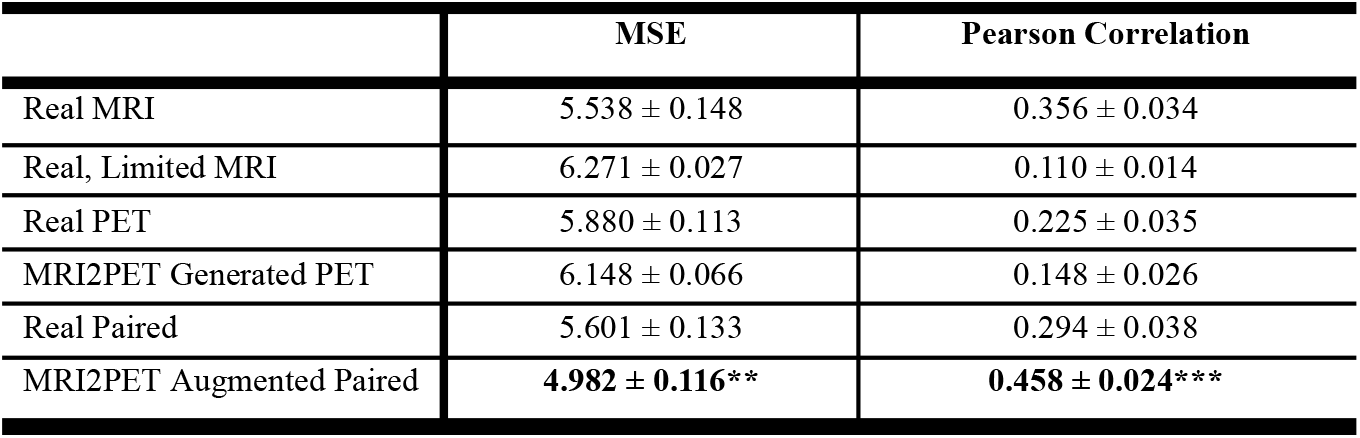
Evaluation results for downstream regression models predicting Mini-Mental State Examination (MMSE) scores. The table presents test performance metrics including Mean Squared Error (MSE) and Pearson Correlation coefficients across different configurations of the original training dataset (real MRI, limited MRI, real PET, synthetic PET, and paired data). Augmented datasets including MRI2PET-generated PET images combined with the limited real paired data yield the lowest error rates and highest correlation, demonstrating substantial improvement in predictive accuracy. Note asterisks refer to p-value significance based on a two-tailed t-test comparing augmented results to the nearest baseline approach with * p < 0.05, **p < 0.01, *** p < 0.001

The synthetic PET dataset alone closely approximated results achieved with real PET data, confirming MRI2PET’s capability to generate clinically informative synthetic scans. Crucially, training with augmented paired datasets resulted in the highest performance improvements, surpassing both real paired and individual modality datasets across accuracy, F1 score, balanced accuracy, AUROC (classification), and mean squared error, and Pearson correlation (MMSE regression).

Specifically, the augmented paired dataset achieved classification accuracy of 0.605 ± 0.007 and F1 scores of 0.595 ± 0.008, representing substantial improvements over all other configurations. Similarly, MMSE prediction with augmented data demonstrated significantly reduced error (4.982 ± 0.116 MSE) and increased correlation (0.458 ± 0.024), clearly outperforming alternative approaches.

These findings underscore MRI2PET’s capability to effectively expand available imaging data, significantly enhancing the robustness, accuracy, and predictive power of downstream machine learning models, demonstrating real-world clinical utility beyond simple data augmentation.

Finally, examining the MMSE predictions offered additional validation. Using a regression model trained exclusively on real PET images, we compared predictions for real and MRI2PET-generated PET scans, revealing a notable positive correlation (r = 0.273). This further affirms MRI2PET’s ability to capture diagnostically relevant information within generated PET scans.

## Discussion

In this study, we introduced MRI2PET, an innovative diffusion-based generative method designed to simulate PET scan imaging from MRI data. MRI2PET specifically addresses significant technical challenges—including the scarcity of paired MRI-PET data, the inherent complexity of 3D volumetric data, and the structural-functional relationships between MRI and PET modalities—by integrating two novel components: a style-transfer-based pre-training strategy and a Laplacian pyramid training objective. Through extensive evaluation using the Alzheimer’s Disease Neuroimaging Initiative (ADNI) dataset, MRI2PET consistently outperformed various generative baselines, demonstrating superior quantitative performance metrics (FID, SSIM, PSNR) and producing qualitatively realistic PET images that closely reflect patient-specific anatomical details.

Beyond the general fidelity and realism of the generated PET images, MRI2PET effectively captured clinically meaningful details, such as brain atrophy and specific patterns associated with Alzheimer’s disease progression. This capability significantly enhanced its practical utility in downstream machine learning applications. Models trained on datasets augmented by synthetic PET images from MRI2PET substantially outperformed those trained solely on real datasets, underscoring the method’s potential to effectively compensate for missing PET imaging data and thereby boost diagnostic and predictive modeling performance.

An intriguing qualitative observation emerged regarding variations in generative image quality across different patient samples. Synthetic PET images generated by MRI2PET exhibited notably greater clarity and anatomical accuracy in patients presenting clear structural brain atrophy on MRI scans, in contrast to cases without evident MRI-detectable decay. Further analysis revealed a consistent pattern: PET images generated for patients at later stages of Alzheimer’s disease (characterized by pronounced atrophy visible on MRI) were significantly more precise and detailed than those at earlier stages without obvious structural changes.

To explain this observation, we proposed a biological hypothesis rooted in the known pathological progression of Alzheimer’s disease, particularly involving amyloid-beta (Aβ) plaque accumulation. Amyloid-beta plaques typically accumulate prior to the structural atrophy visible through MRI, which is itself driven by neuronal loss and tau disease pathology. Thus, patients at early disease stages—who might exhibit substantial underlying amyloid pathology yet no visible structural MRI abnormalities—can present multiple plausible metabolic patterns detectable by PET. The generative uncertainty associated with these multiple potential outcomes may lead MRI2PET to produce less distinct, blurred synthetic images as the model implicitly averages across these scenarios rather than clearly delineating one distinct outcome.

Although this averaging behavior is suboptimal from a purely generative modeling perspective, where distinct scenario generation would be preferable, it significantly highlights MRI2PET’s responsiveness to underlying biological processes. Indeed, this phenomenon suggests that MRI2PET is implicitly sensitive to biologically meaningful variations related to early Alzheimer’s pathology that structural MRI alone cannot reveal. Recognizing this biological underpinning provides valuable insights into possible avenues for future model refinement, such as explicitly modeling uncertainty or incorporating early-stage biomarkers.

Therefore, while the observed variability in generative quality indicates opportunities for technical improvement, it also underscores MRI2PET’s potential utility as a tool for capturing subtle pathological insights, potentially facilitating earlier diagnosis and more precise differentiation among various Alzheimer’s disease stages.

It also underscores important limitations to be aware of within the core task setup. Generating PET images based on MRI scan results can by definition inject no additional information than is already available within that MRI. This does not reduce the value or potential applications. There may be lots of information that is not readily discerned from looking at the MRI itself, we have shown in this paper that we succeed in injecting utility through the augmentation of MRI-PET datasets, and the ability to simulate a specific instantiation within the range of potential corresponding outcomes, especially a physiologically meaningful one, offers clearly valuable applications. However, it does mean that this value and these applications should be conditioned and this setup should be noted.

In summary, MRI2PET represents a promising and clinically relevant advancement toward simulating PET imaging outcomes from MRI data, delivering immediate practical utility and opening avenues for further methodological refinements and enhanced biological interpretability.

## Data Availability

The ADNI,^22^ UKBiobank,^37^ and PPMI ^20^ that we use are publicly available and may be downloaded and used freely after applying at and https://ida.loni.usc.edu/login.jsp and https://ams.ukbiobank.ac.uk/ams/.

## Code Availability

All original code has been deposited at Zenodo under the DOI 10.5281/zenodo.15089724 and is publicly available there as well as at https://github.com/btheodorou99/MRI2PET/.

## Acknowledgements

This work was supported by NSF award SCH-2205289, SCH-2014438, and IIS-2034479. This research was also supported in part by the Intramural Research Program of the NIH, National Institute on Aging (NIA), National Institutes of Health, Department of Health and Human Services; project number ZO1 AG000534, as well as the National Institute of Neurological Disorders and Stroke (NINDS).

## Author Contributions

F.F. designed and oversaw the study. B.T., A.D., B.A., J.S., and F.F. did the primary interpretation of the data. B.T. conducted all the experiments. B.T., J.S., and F.F. wrote the manuscript. A.D., B.A., and M.N. made major contributions to manuscript editing. B.T., A.D., B.A., J.S., and F.F. verified the data. All authors had access to all the data in the study and had final responsibility for the decision to submit for publication.

## Data Sharing

Code for preprocessing the data, training all compared methods, and generating all results is available online at https://github.com/btheodorou99/MRI2PET. Each of the ADNI, PPMI, and UKBiobank datasets are available upon application at https://adni.loni.usc.edu/data-samples/adni-data/, https://www.ppmi-info.org/access-data-specimens/, and https://www.ukbiobank.ac.uk/ respectively. For further information please contact.

