## Supplemental Material for "MRI2PET: Realistic PET Image Synthesis from MRI for Automated Inference of Brain Atrophy and Alzheimer’s"

### Supplementary Material

#### Experimental Setup

To conduct our experiments, we train each compared method and generate PET scan images based on each MRI scan in our test dataset. We also generate PET results based on all MRI images from the ADNI dataset in our other splits with just our MRI2PET model. We then perform a variety of analyses comparing the similarity in terms of quantitative correspondence, visual adherence, and downstream utility of these generated PET scans to the true PET results. We report 95% confidence intervals for each experimental value for each metric, calculated by performing 100 bootstraps of the test set. We present the best values in bold for each metric, and we bold any other values whose means fall into that best value's confidence interval. We perform all experimentation using Nvidia A100 GPUs on the National Institutes of Health's Biowulf cluster (see https://hpc.nih.gov for reference). All source code for our experiments can be found at https://github.com/btheodorou99/MRI2PET/.

#### Dataset Statistics

To provide more detailed information regarding our datasets and offer context for our experiments, we provide more detailed count and demographic information in a pair of tables. First, we provide a count of where the MRI images were found across our three datasets in Supplementary Table 1. Then, we provide a variety of demographics and counts, especially pertaining to our downstream experiments and their labels in Supplementary Table 2.

| Total | ADNI | PPMI | UKBiobank |
|---|---|---|---|
| 22,956 | 11,453 | 1,956 | 9,547 |

Supplementary Table 1: Number of MRI Images by dataset

| | Train | Validation | Test |
|---|---|---|---|
| Number of PET Scans | 2,017 | 225 | 250 |
| Number of Patients | 1,068 | 205 | 232 |
| Percent Male | 51.2% | 55.9% | 48.7% |
| Average Age at Intake | 71.93 ± 7.12 | 71.36 ± 6.40 | 71.06 ± 6.66 |
| Number of CN | 393 | 89 | 98 |
| Number of MCI | 355 | 63 | 67 |
| Number of AD | 306 | 52 | 66 |
| Average MMSE | 25.75 ± 5.03 | 26.37 ± 4.55 | 25.62 ± 5.56 |
| Number MMSE 20+ | 905 | 182 | 195 |
| Average MMSE 20+ | 27.16 ± 2.75 | 27.46 ± 2.45 | 27.28 ± 2.76 |

Supplementary Table 2: Dataset Statistics and Demographics of the ADNI PET Dataset

#### Problem Formulation

We begin the presentation of our method by introducing the data, problem, and task using notation that we will build upon in describing our approach.

***Definition 1*** *(MRI Scan Imaging).* We denote the MRI scan data as $M \in R^{d_m \times r_m \times r_m}$ where $d_m$ represents the depth or number of slices and $r_m$ represents the height and width resolution of the image. Each pixel or variable in $M$ is normalized to lie within a defined range (which we set to be [0, 1]).

***Definition 2*** *(PET Scan Imaging).* We then similarly represent the PET scan data as $P \in R^{d_p \times r_p \times r_p}$ where $d_p$ is the PET-specific depth and $r_p$ is the PET-specific resolution of the image. $P$ is normalized to lie within a standard normal distribution for use in diffusion models, though it is then converted to the range [0, 1] for visualization.

***Task 3*** *(MRI to PET Generation).* The task is then, given an MRI scan $M$, to generate the corresponding PET scan $P$ for the same patient such that it closely mirrors the true PET results. This is achieved by learning and sampling from $P(P \mid M)$.

#### 3D Diffusion Framework

We utilize a standard diffusion process to transform a sample from a simple source distribution of Gaussian noise to one from a more complex target distribution of PET images via a series of small diffusive adjustments.

**Iterative Diffusion** The forward noising pathway consists of repeatedly adding noise to an initial PET image until it resembles a fully noisy sample from our source Gaussian distribution. The PET image is then recovered by reversing the process and denoising the noisy sample step-by-step. Formally, the noising process at diffusion step $t$ is represented by:

$$d\hat{\mathcal{P}}^{(t)} = \sqrt{\hat{\alpha}_t}\hat{\mathcal{P}}^{(t)}dt + \sqrt{1-\hat{\alpha}_t}dW^{(t)}$$

Where $\hat{\mathcal{P}}^{(t)}$ denotes the noisy PET image at step $t$, $\sqrt{\alpha_{\hat{t}}}$ is the drift term that scales the image intensity, $\sqrt{1-\alpha_{\hat{t}}}$ is the diffusion term that scales the noise, and $dW$ is an infinitesimal Wiener process representing the random noise $\varepsilon$. We discretize this continuous diffusion over $T = 1000$ noise steps, and we set $\hat{\alpha}_t$ by defining $\beta$ as a 1000-step linear spacing between $\beta_{\text{start}} = 0.0015$ and $\beta_{\text{end}} = 0.02$ with $\alpha = 1 - \beta$ and $\hat{\alpha}_t$ is finally the cumulative product of the first $t$ values of $\alpha$.

**Denoising with DDPM** We conduct the reverse denoising process using a neural network which serves as our denoising diffusion probabilistic model (DDPM). This model directs the denoising process at each diffusion step given the noisy image and conditioning information (in our case, the time step number and MRI image). It does this by predicting the error $\epsilon$ between the provided noisy image $\hat{\mathcal{P}}^{(t)}$ image and the true target image $\mathcal{P}^{(t)}$. The generation process then takes an input image initialized as pure Gaussian noise and iteratively feeds that image along with the corresponding MRI to the trained model to repeatedly refine the output and generate realistic PET scan results.

We use a UNet autoencoder as our DDPM as is standard within the domain for use with diffusion models.[32] UNet architectures consist of an encoding pathway, which embeds the input image into lower-dimensional space, and a subsequent decoding pathway which maps that embedding back into the original dimensionality. Each step in either pathway is furthermore connected via a skip connection to maintain spatial information as well as the relevant conditioning information. The only adaptations we make onto the architecture here are replacing the standard 2D convolutions with 3D versions (representing our PET images as a single channel 3D structure rather than a multi-channel 2D structure) and replacing the standard complete attention with a linear version to reduce the computational complexity to accommodate our computing resources.

#### Laplacian Pyramid Loss Formalization

We add an additional loss component onto the standard mean squared error-based training objective which instructs the DDPM to accurately predict the noise. This additional objective aims to enhance the model's ability to capture important details at multiple resolutions within the generated images. It does so by leveraging the Laplacian pyramid, a multi-scale image representation consisting of a set of residuals between increasingly blurred and lower-dimensional images.

Specifically, a Laplacian pyramid is constructed by first creating a Gaussian pyramid by iteratively downsampling (through a 2x2 averaging pooling operation) the original image. The Laplacian pyramid then takes each layer of the Gaussian pyramid and compares it to the layer below, upsampled via bilinear interpolation back to the same size. So, it effectively compares the original image, scaled to different, increasingly small, resolutions, with blurrier versions of itself. The differences in these comparisons are interpreted to be the details at a given resolution.

We can formalize the conceptual Laplacian Pyramid loss calculation introduced in our main paper as

$$\mathcal{L}_{\text{Lap}}(\mathcal{P}, \hat{\mathcal{P}}) = \sum_{l=1}^{5} \text{MSE}(\text{LapPyr}(\mathcal{P})_l, \text{LapPyr}(\hat{\mathcal{P}})_l)$$

where a given Laplacian Pyramid is calculated by

$$\text{LapPyr}(P) = \{P_l - \text{Upsample}(\text{Downsample}(P_l))\}_{l=1}^{5}$$

and the total loss function for training the DDPM is then defined as a weighted combination of the standard noise prediction loss and this Laplacian pyramid loss:

$$\mathcal{L} = \text{MSE}(\epsilon, \tilde{\epsilon}) + \lambda \mathcal{L}_{\text{Lap}}(\mathcal{P}, \tilde{\mathcal{P}})$$

where $\epsilon$ is the noise added to $\mathcal{P}$ to produce the model input $\hat{\mathcal{P}}$, $\tilde{\epsilon}$ is the DDPM model's noise prediction, $\tilde{\mathcal{P}}$ is the predicted image based on that predicted noise as calculated by $\tilde{\mathcal{P}} = \frac{1}{\sqrt{\hat{\alpha}}} \cdot \hat{\mathcal{P}} - \frac{\sqrt{1-\hat{\alpha}}}{\hat{\alpha}} \cdot \tilde{\epsilon}$ and $\lambda$ is the weighting factor, which we experimentally set to 0.25.
